# SARS-CoV-2 surveillance in decedents in a large, urban medical examiner’s office

**DOI:** 10.1101/2020.08.03.20162883

**Authors:** Andrew F. Brouwer, Jeffrey L. Myers, Emily T. Martin, Kristine E. Konopka, Adam S. Lauring, Marisa C. Eisenberg, Paul R. Lephart, Teresa Nguyen, Andrea Jaworski, Carl J. Schmidt

## Abstract

**Background:** SARS-CoV-2 has become a global pandemic. Given the challenges in implementing widespread SARS-CoV-2 testing, there is increasing interest in alternative surveillance strategies.

**Methods:** We tested nasopharyngeal swabs from 821 decedents in the Wayne County Medical Examiner’s office for SARS-CoV-2. All decedents were assessed by a COVID-19 checklist, and decedents flagged by the checklist (237) were preferentially tested. A random sample of decedents not flagged by the checklist were also tested (584). We statistically analyzed the characteristics of decedents (age, sex, race, and manner of death), differentiating between those flagged by the checklist and not and between those SARS-CoV-2 positive and not.

**Results:** Decedents were more likely to be male (70% vs 48%) and Black (55% vs 36%) than the catchment population. Seven-day average percent positivity among flagged decedents closely matched the trajectory of percent positivity in the catchment population, particularly during the peak of the outbreak (March and April). After a lull in May to mid-June, new positive tests in late June coincided with increased case detection in the catchment. We found large racial disparities in test results: despite no statistical difference in the racial distribution between those flagged and not, SARS-CoV-2 positive decedents were substantially more likely to be Black (89% vs 51%). SARS-CoV-2 positive decedents were also more likely to be older and to have died of natural causes, including of COVID-19 disease.

**Conclusions:** Disease surveillance through medical examiners and coroners could supplement other forms of surveillance and may serve as a possible early outbreak warning sign.

## Introduction

COVID-19, the disease caused by coronavirus SAR-CoV-2, has become a global pandemic, with more than 2.9 million cases and 130,000 deaths reported in the United States [1] and 11.5 million cases and 0.5 million deaths reported globally [2]. Public health surveillance and detection of cases has been challenging, particularly in the early months of the outbreak, as tests and supplies for this novel pathogen needed to be developed, validated, produced, and distributed. Although widespread testing of the population is likely the most accurate surveillance strategy, it is also logistically challenging and costly. Consequently, there is increasing interest in alternative surveillance strategies. Tracking reports of coronavirus-like illness (CLI), akin to the tracking of influenza-like illness (ILI), is one supplemental strategy that, while relatively easy to implement, lacks specificity and must be coupled with lab-confirmed surveillance when multiple respiratory pathogens are circulating [3]. Environmental surveillance of SARS-CoV-2 in wastewater also shows some promise but may be difficult to implement widely and may be difficult to interpret in terms of actual numbers of infected people [4-6].

Disease surveillance through testing decedents in medical examiner’s and coroner’s offices may offer a supplemental perspective on the outbreak, particularly as it can detect the virus in those that were not clinically diagnosed when alive. The Wayne County Medical Examiner (WCME) provides pathology services, including autopsies, for Wayne and Monroe Counties in Michigan, including the city of Detroit. This office has a catchment of approximately 1.9 million people. Since mid-March (shortly after surveillance networks began detecting positive cases [7]), WCME has been piloting daily SARS-CoV-2 surveillance by testing nasopharyngeal swabs of decedents, including both COVID-19 suspects and non-suspects. In this analysis we compare percent positivity in WCME’s piloted SARS-Cov-2 surveillance among decedents—distinguishing between those flagged by a COVID-19 checklist and those that were not—to the percent positivity of tests among people in the surrounding catchment area. We propose that decedent surveillance may offer a cost-effective supplemental surveillance strategy for SARS-CoV-2 and other pathogens.

## Methods

The Wayne County Medical Examiner serves Wayne and Monroe Counties in Michigan, including the city of Detroit. The catchment of this office was 1,899,843 people in 2019 [8]. Starting March 16, 2020, WCMEO tested nasopharyngeal swabs from up to 10 decedents per day for SARS-CoV-2. A total of 821 decedents were tested through July 10, 2020. Cases with significant head trauma, anterior lividity, or signs of decomposition were excluded from the pool of possible testing because of previous experience suggesting that most swabs on these cases would return invalid results. Decedents were assessed for possible recent COVID-19 illness through a checklist (presumptive or confirmed COVID-19 diagnosis; signs of infection (fever, shortness of breath, sneezing, coughing, chest pain, body aches); recent travel; contacts, family, or friends with suspected or confirmed COVID-19 diagnosis or signs of infection), and flagged decedents (i.e., those with one or more of the listed items) were swabbed preferentially. Decedents that were not flagged were tested at random. Specimens were tested using either the RealTime m2000 SARS-CoV-2 Assay (Abbott Molecular, Des Plaines, IL) or Simplexa COVID-19 Direct Kit (DiaSorin, Cypress, CA).

Data on percent positivity of SARS-CoV-2 tests administered in people in the WCMEO catchment area were obtained from the Michigan Department of Health and Human Services through the MI Start Map dashboard (https://www.mistartmap.info/) [9].

We determined the characteristics of the decedent population, specifically age (not available for 2 decedents); sex (female, male); race (Black, White, or Other/Unknown); manner of death (natural (cardiovascular), natural (other), accident, homicide/suicide, or pending/indeterminant); and SARS-CoV-2 status (negative, positive, invalid/inconclusive). We compared the characteristics of the decedents that were vs those that were not flagged by the COVID-19 checklist; we also compared the characteristics of decedents that were vs those that were not SARS-CoV-2 positive. Comparisons between groups were assessed by t-test (age), test of proportions (sex), or chi-square test (race (only among Black and White participants due to low numbers of other decedents of other or unknown race), manner of death, and SARS-CoV-2 status).

## Results

The characteristics of the tested decedent population—overall and distinguished by COVID-19 checklist flag and test result—are given in Table 1. The average age at death was 46 years. There were significant differences in the age distribution between both decedents that were and were not flagged for testing and those that did and did not test positive for SARS-CoV-2. A larger fraction of those flagged by the checklist were older: 80% of decedents who were flagged were over 40 years old, compared to 59% of those who were not flagged. Similarly, a larger fraction of those positive for SARS-CoV-2 were older: 80% of those that tested positive were over 40 years old, compared to only 64% of those that tested negative. There were no significant differences by sex, though decedents were overall more likely to be male (70%) than the catchment population (48% [8]).

**Table 1:**
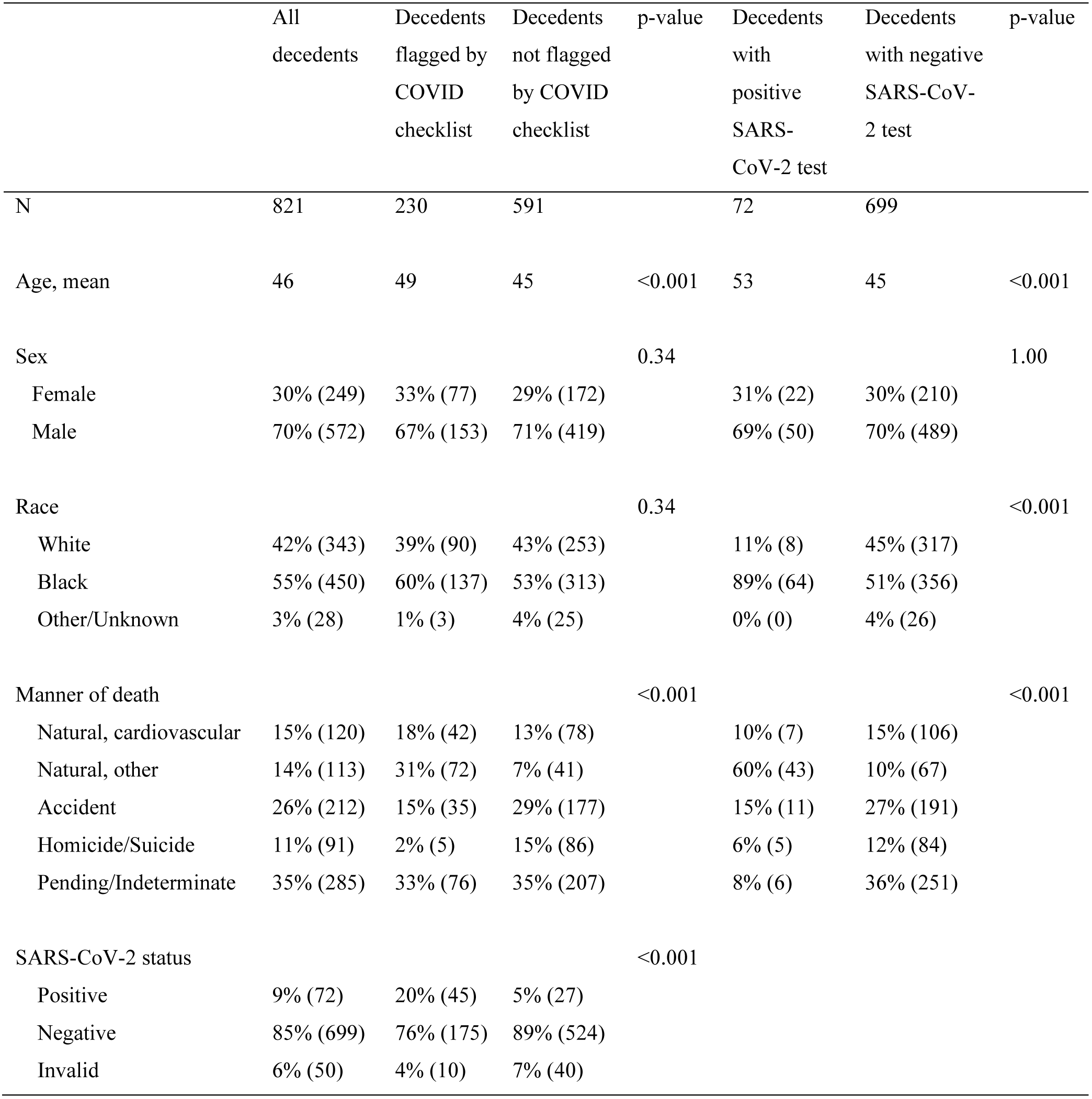
Characteristics of the decedent population tested for SARS-CoV-2 in the Wayne County Medical Examiner’s Office.

The proportion of decedents that were Black (55%) was greater than in the catchment (36% [8]). There were no racial differences between the groups that were or were not flagged. However, there were stark racial differences between the group of decedents that tested positive for SARS-CoV-2 and the group that tested negative; 89% of those that tested positive were Black, while only 51% of those that tested negative were Black. This disparity was more pronounced among SARS-CoV-2 positive decedents that were not flagged by the checklist (96% Black vs. 4% White) than among those that were flagged (85% Black vs 15% White).

At the time of submission, about 65% of decedents had a determined cause of death. There were a significantly higher percentage of deaths determined to be natural among both those that were flagged and those that tested positive. There was also a lower fraction of cases without a cause of death determination among the SARS-CoV-2 positive decedents because most positive tests were from the early phase of the outbreak. The flagged decedents did have a significantly higher percentage of positive SARS-CoV-2 tests (18% vs. 5%).

We present the seven-day average percent positivity for all decedents, flagged decedents, and decedents that were not flagged in Figure 1. We also include the seven-day average percent positivity among all tests in people in the catchment for comparison. Percent positivity changed substantially over time, peaking at the end of March. The percent positivity among flagged decedents closely matches the percent positivity in the catchment population. The percent positivity among the decedents that were not flagged also followed a similar trajectory but was lower overall.

**Figure 1:**
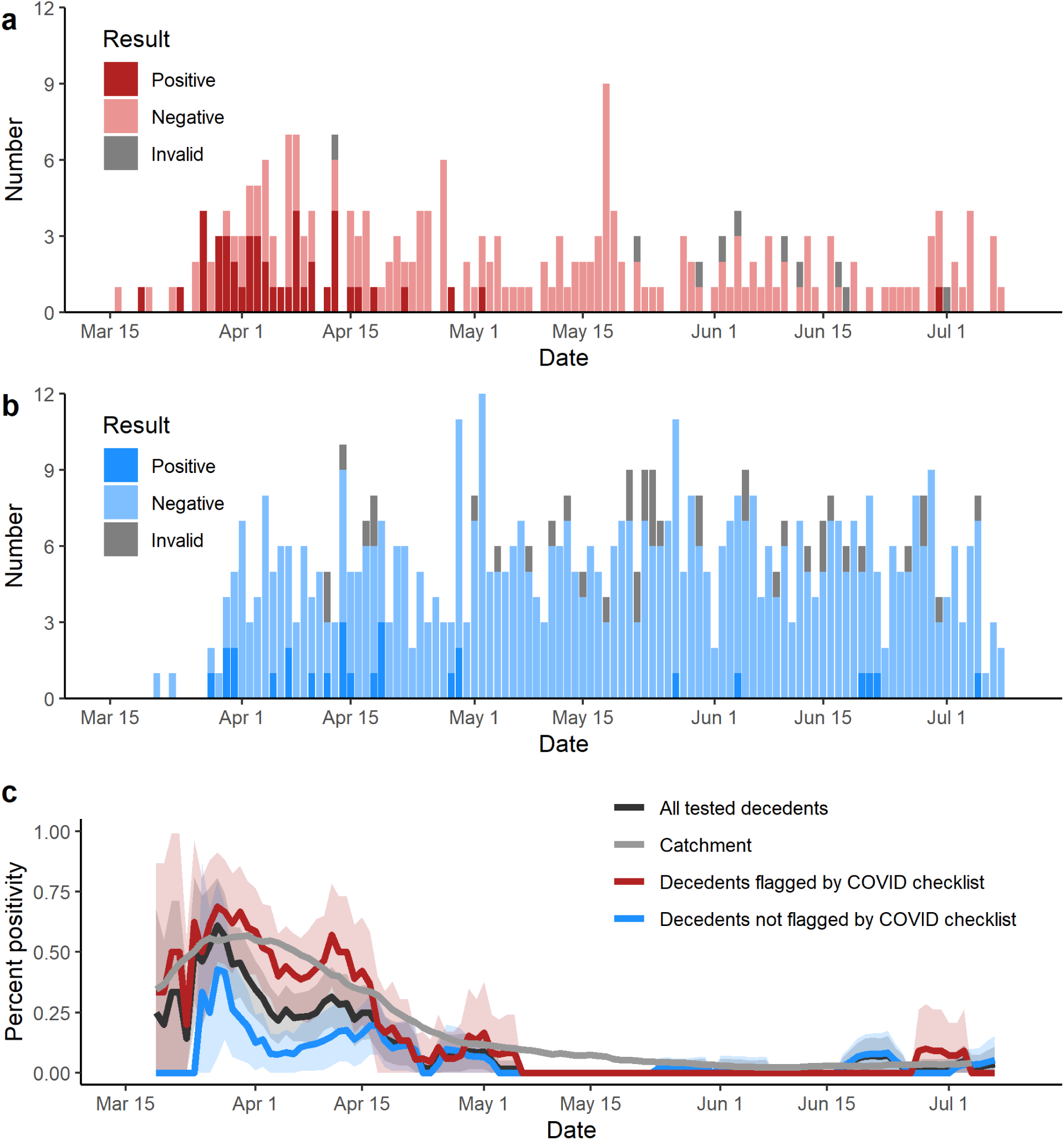
a) SARS-CoV-2 test results among decedents flagged by the COVID-19 checklist. b) SARS-CoV-2 test results among decedents not flagged by the COVID-19 checklist. c) Seven-day average percent positivity in decedents and in the catchment population; we further distinguish between decedents that were flagged by the COVID-19 checklist and those that were not.

## Discussion

Our work demonstrates that disease surveillance through medical examiners and coroners could offer a cost-effective supplemental form of disease surveillance in urban areas and may serve as possibly early warning sign. The percent positivity for SARS-CoV-2 infection among decedents flagged for testing by a COVID-19 checklist in large, urban medical examiner’s office closely mirrored percent positivity among tests in the catchment population. The percent positivity among decedents that were not flagged followed a similar trajectory but with a lower magnitude; testing decedents that are not COVID-19 suspects may improve our understanding of asymptomatic or atypical disease presentation. Few cases were detected between early May and mid-June, which was a time of low case incidence in the catchment, but new decedent cases were detected as case incidence began to increase again in late June [9]. More work is needed to refine sampling protocols and expand capacity to maximize the accuracy and effectiveness of this surveillance method.

In this analysis, we found significant differences in the characteristics of decedents that were or were not flagged by the COVID-19 checklist. Flagged decedents were more likely to be older, have died of natural causes, and to have tested positive for SARS-CoV-2. Each of these differences might be anticipated given the goal of the COVID-19 checklist and the profile of patients with symptomatic COVID-19 disease. It is unclear whether the 5% positivity among decedents that were not flagged represent asymptomatic/presymptomatic infections in those that died of unrelated causes or COVID-19 related deaths with atypical presentation. Future work will be needed to better understand these positives, but the diverse nature of COVID-19 symptoms [10] increases the likelihood of the latter explanation and underscores the danger in overreliance on case definitions for emerging infection surveillance. It has been suggested that case definitions may be underestimating the relevance of COVID-19 to increased likelihood of stroke or other cardiovascular death [11-14]. This study was not designed to address this question directly, but we did find a small fraction (10%) of SARS-CoV-2 positive patients assigned a cardiovascular cause of death; all had been flagged by the COVID-19 checklist.

We also found significant difference in the characteristics of decedents that were or were not positive for SARS-CoV-2. Positive decedents were more likely to be older, to be black (89% vs 51%), and to have died of natural causes than negative decedents. The difference in SARS-CoV-2 positivity by race is alarming, particularly as there was no difference in the racial distribution of those that were flagged by the COVID-19 checklist. The disparity was even more pronounced among those positives not flagged by the checklist. These findings underscore the severe health disparities for Black people in this pandemic (and more generally) in the U.S. [15]. As recent perspectives point out, we must understand these disparities in the context of racism, socioeconomic disparities, enhanced susceptibility due to chronic stress, prevalence of underlying risk factors, and geography of transmission [16-18].

Epidemiological surveillance based on data from medical examiners is naturally used for certain kinds of cause-of-death surveillance and is used in some national databases, such as the National Violent Death Reporting Systems [19]. However, there are general limitations to the use of medical examiner data for epidemiological surveillance [20-22]. First, the population of decedents entering a medical examiner’s office may not be representative of the catchment population. Here, a higher percentage of decedents were male and black than the catchment population, reflecting entrenched institutional and sociocultural pressures resulting in a higher likelihood of a cause of death that would fall within the medical examiner’s purview. Any surveillance program will need to recognize the limitations caused by the skewed population. A second limitation of surveillance from medical examiner’s offices is that autopsies are apt to reveal medical conditions that would not have been detected while the decedent was alive. From the point of view of SARS-CoV-2 surveillance, detection of infections that were not clinically detected during life offers an opportunity to better understand atypical or asymptomatic presentations of the disease (particularly when the cause of death is unrelated to the infection) and to detect increased community transmission prior to large increases in symptomatic cases. Because of the limitations of decedent surveillance, it has largely been used only in small, targeted studies, such as those looking for underreporting of disease related to occupational exposure (e.g., silicosis [23]). Arguably, the potential of decedent disease surveillance has been underappreciated. While its limitations make it a poor choice for surveillance of many diseases, there may be value in expanding decedent testing around epidemic pathogens [24]—not only SARS-CoV-2 but perhaps also influenza or other, similar diseases that tend to be widespread.

The results from a pilot surveillance program for SARS-CoV-2 in a large, urban medical examiner’s office suggest that expanded SARS-CoV-2 testing in medical examiners’ and coroners’ offices may offer an additional surveillance stream to support outbreak response. In our analysis, percent positivity among decedents flagged by a COVID-19 checklist closely matched that of the catchment population. This study also revealed large racial health disparities in SARS-CoV-2 infection among decedents.

## Data Availability

Relevant data will be made available once the manuscript has been accepted in a peer-reviewed journal.

## Acknowledgments

Data collection was funded by the Wayne County Medical Examiner through the office’s normal operating budget.

